# Forecasting trachoma control and identifying transmission-hotspots

**DOI:** 10.1101/2020.12.21.20248631

**Authors:** Seth Blumberg, Joaquin M Prada, Christine Tedijanto, Michael S Deiner, William W. Godwin, Paul M. Emerson, Pamela J Hooper, Anna Borlase, T. Deirdre Hollingsworth, Catherine E. Oldenburg, Travis C Porco, Benjamin F. Arnold, Thomas M Lietman, Trachoma survey collaborators

## Abstract

**Background:** Tremendous progress towards elimination of trachoma as a public health problem has been made. However, there are areas where the clinical indicator of disease, trachomatous inflammation—follicular (TF), remains prevalent. We quantify the progress that has been made, and forecast how TF prevalence will evolve with current interventions. We also determine the probability that a district is a transmission-hotspot based on its TF prevalence (i.e. reproduction number greater than one).

**Methods:** Data on trachoma prevalence comes from the GET2020 global repository organized by the World Health Organization and the International Trachoma Initiative. Forecasts of TF prevalence and the percent of districts achieving local control is achieved by regressing the coefficients of a fitted exponential distribution for the year-by-year distribution of TF prevalence. The probability of a district being a transmission-hotspot is extrapolated from the residuals of the regression.

**Results:** Forecasts suggest that with current interventions, 96.5% of surveyed districts will have TF prevalence among children aged 1-9 years <5% by 2030 (95% CI: 86.6-100.0%). Districts with TF prevalence < 20% appear unlikely to be transmission-hotspots. However, a district having TF prevalence of over 28% in 2016-2019 corresponds to at least 50% probability of being a transmission-hotspot.

**Conclusions:** Sustainable control of trachoma appears achievable. However there are transmission-hotspots that are not responding to annual mass drug administration of azithromycin and require enhanced treatment in order to reach local control

## Introduction

Although trachoma has caused irreversible blindness or visual impairment in 1.9 million people in 44 countries, elimination as a public health problem (henceforth referred to as trachoma ‘control’) is within reach.^1,2^ The World Health Organization Alliance for the Global Elimination of Trachoma by the year 2020 (GET2020) has coordinated a combination of approaches including surgery, mass drug administration (MDA) of antibiotics, promotion of facial cleanliness, and environmental improvement to reduce the burden of trachoma.^3,4^ Progress is monitored by clinical surveys that measure the prevalence of trachomatous inflammation—follicular (TF) in individual health districts.^5–7^ An azithromycin donation program to facilitate antibiotic MDA began operations in 1999. Since then, many districts have shown marked reductions in the prevalence of trachoma.^8^ Of 48 countries previously identified as having trachoma, ten countries have now been validated by the World Health Organization (WHO) as having achieved control.^4,9^ However there are some districts with sustained transmission in the remaining 38 countries that require additional intervention in order for control to be achieved.^1^

A challenge in conducting trachoma surveillance is that the prevalence of the causative bacterium, *Chlamydia trachomatis*, the bacterial load, and the transmission intensity, are all difficult to measure. The WHO-recommended cross-sectional surveys of TF prevalence only measure a downstream inflammatory complication of actual infection. However, monitoring the trend in TF prevalence over time provides information about the trajectory of disease and hence trachoma transmission.^10,11^ In addition, recent studies have indicated that a signature of self-limited ‘subcritical’ disease transmission (i.e. reproduction number is less than one) is that the distribution of district-level TF measurements worldwide converges to an exponential distribution.^12,13^ Evidence of subcritical transmission is important, since that would imply that current interventions will lead to global control.^14^

A second challenge for surveillance is to identify transmission-hotspots, which we define as districts that demonstrate a reproduction number greater than one (i.e. sustained transmission), despite ongoing control programs. These districts might benefit from MDA with azithromycin that is more frequent than the annual cycle routinely used by country programs.^15–17^ Traditionally, hotspot districts have been classified based solely on the current burden of TF.^18–20^ However, since TF prevalence is not a direct measure of transmission, hotspots identified solely by TF prevalence may or may not be a transmission-hotspot. To reconcile these two approaches, a single measurement of TF prevalence can be viewed as offering a probability that a particular district has sustained transmission.

Here we use the GET2020 database of district-level prevalence estimates to address the aforementioned challenges. In particular the distribution of TF prevalence across districts is used to assess the trends in disease transmission, and whether the data are consistent with progress towards global elimination. These results are then used to forecast how progress towards global control is expected to change over time. Lastly, the probability of a district being a transmission-hotspot is estimated as a function of TF prevalence.

## Methods

### Data

Data on TF prevalence were obtained from the GET2020 database, which serves as a major tool in assessing progress towards elimination. The dataset consists of time-stamped estimates of TF prevalence from individual health districts. Since each datum represents aggregation over a large geographical area and typically represents a population of 100,000-250,000 individuals, analysis was deemed exempt from review by the University of California institutional review board. Each TF prevalence measurement is limited to clinical examination of children aged one to nine.

### Modeling approach

We utilized a susceptible-infected-susceptible model for trachoma transmission.^14^ To address the temporal dynamics of TF prevalence, the observed TF prevalence distribution for each year was modelled as an exponential distribution with a rate parameter that decays exponentially with time. Our model was calibrated by maximizing the log-likelihood of the entire data set. The temporal dynamics of these parameters were then extrapolated to produce a nowcast for 2020 and forecasts for 2021-2030. Confidence intervals for our analyses were determined by bootstrapping at the country level. For a specific observed value of TF, we estimate the probability that a district is a transmission-hotspot by quantifying how much of the observed probability density exceeds the best-fit of an exponential distribution that would be representative of entirely subcritical transmission.

More details about the data and modeling approach can be found in the supplementary material (Text S1).

## Results

### Surveillance

After excluding duplicate entries, surveys that had an undefined survey type, and surveys with TF prevalence < 0.5 percent, there were 3588 prevalence measurements in the GET2020 data for 2004-2019. The data represented 52 countries and 1621 unique districts.

The distributions of TF prevalence show the number of districts surveyed has increased substantially from 2004 to 2019 (Figure 1). Also, the proportion of districts with TF less than 5% has increased over time. Finally, the shape of the prevalence distribution has evolved from multi-modal distribution into a uniformly decreasing distribution, akin to an exponential distribution.

**Figure 1.**
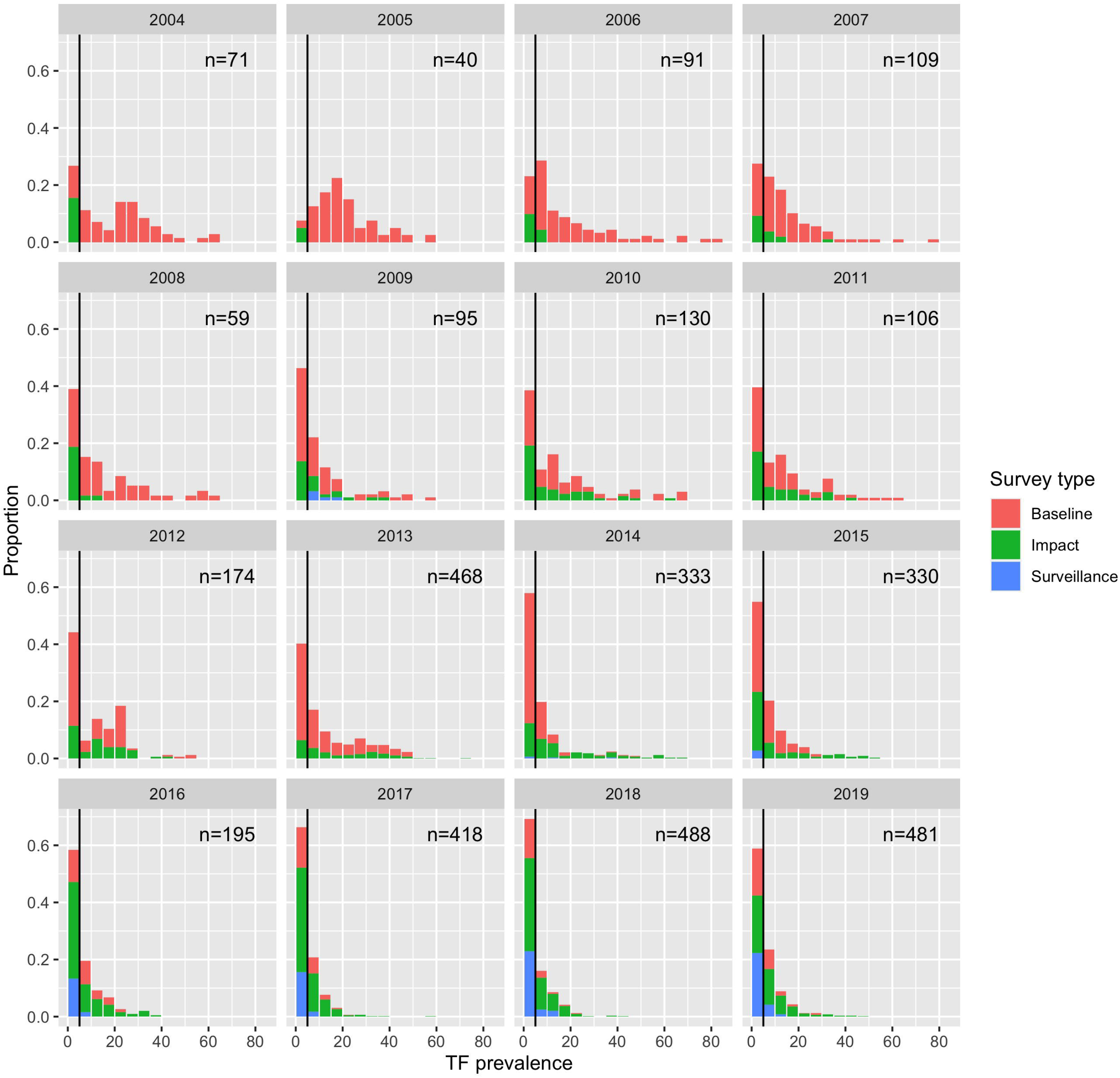
Distribution of district-level prevalence of TF as recorded by the GET2020 Alliance. Each panel represents a different year of data collection. Bin heights represent the number of health districts sampled that had a TF prevalence within the five percent range of each bin. The black line indicates the WHO TF < 5% threshold for district-level control. Colors represent the type of survey with baseline occurring before MDA, impact occurring soon after MDA, and surveillance used to confirm local control.

Stratification of TF prevalence by survey type shows additional insight (Figures 1 and S1). Baseline, impact and surveillance survey all show a large proportion of surveys with TF < 5% in 2016-2019. Meanwhile the number of baseline surveys is decreasing as most areas with concern for trachoma have now been surveyed. Meanwhile, the number of impact and surveillance surveys are increasing. Thus the decrease in the leftward shift of the overall TF distribution likely reflects the impact of annual MDA.

### Forecasts

We utilized probabilistic regression on the GET2020 data to provide a statistical forecast for the distribution of TF in 2020 and beyond (Figure 2). Even with the substantial confidence intervals introduced by bootstrapping at the country level, clear progress towards trachoma control can be seen in the temporal decrease of mean TF prevalence, and the increase in the number of districts that have achieved a TF prevalence of less than five percent from 2004-2019.

**Figure 2.**
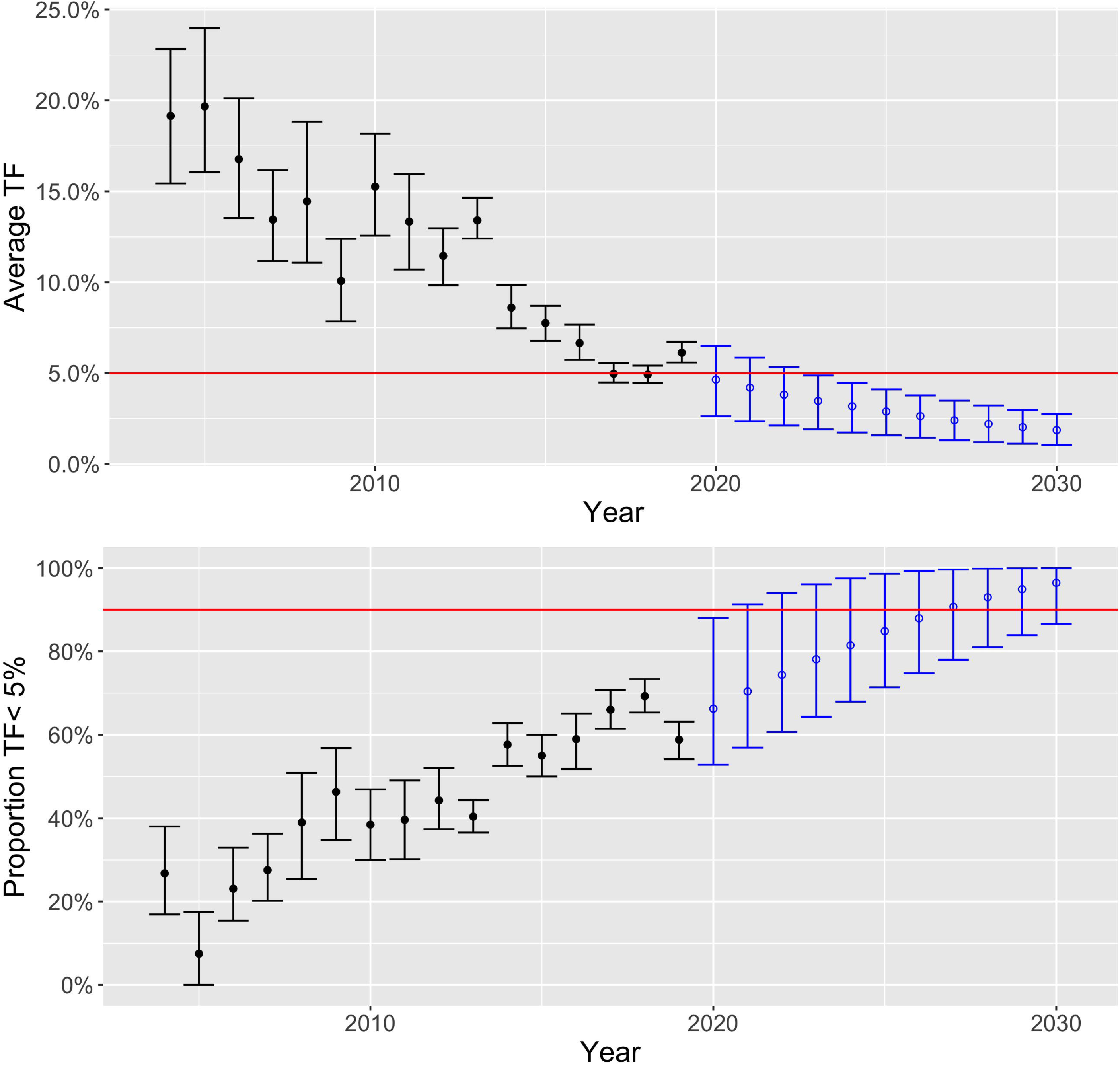
Mean TF prevalence of all surveyed districts (top) and percent of surveyed districts with TF less than 5 percent (bottom). Median estimates and 95 percent confidence intervals are shown, as determined by bootstrap analysis. Error bars with closed circles represent results from sampling retrospective data with replacement. The error bars with open circles are based on probabilistic forecasts. The horizontal lines indicate an average TF of 5% (top) and a control target of having 90% of districts with TF < 5% (bottom).

Our forecast using an exponential regression model suggests that with 98.5% certainty the average TF prevalence across the districts in the GET2020 database will be below 5% by 2023 (Figure S2 and 2 - top). Furthermore by 2030, our model estimated that there is a 97.5% chance that at least 86.6% of districts would have a TF prevalence below 5% (Figure 2 - bottom).

Results for the other model distributions considered were either quite similar to the exponential regression forecast (e.g. gamma regression in FIgure S3) or became unstable for long-range forecasts (beta and lomax regression in FIgure S3). Our ensemble of bootstraps estimates the overall *R* to be 0.95 (95% CI: 0.94-0.96) for 2020.

Although the Global Trachoma Programme has not met its elimination goals by 2020, there is clear evidence of longitudinal success. For concreteness we define a ‘control target’ as occurring when over 90% of the sampled districts have TF less than 5% (excluding district surveys with TF < 0.5% as these may never have had endemic trachoma). With this definition, our forecast anticipates a probability of 35% and 90% that control will be achieved by 2025, and 2030 respectively (Figure S2).

### Transmission-hotspots

Our estimated probability of being a transmission-hotspot increases as TF prevalence increases (Figure 3). In addition, the TF prevalence at which there is >50% probability of supercritical transmission decreases over time (seen by the left shift of 2016-2019 in Figure 3). This could be because the widespread distribution of MDA has increasingly suppressed those districts that may have initially had relatively high TF prevalence, but are not transmission-hotspots since they respond to MDA. Districts with TF < 20% likely represent subcritical transmission headed towards local elimination, even though disease may not yet be locally controlled.

**Figure 3.**
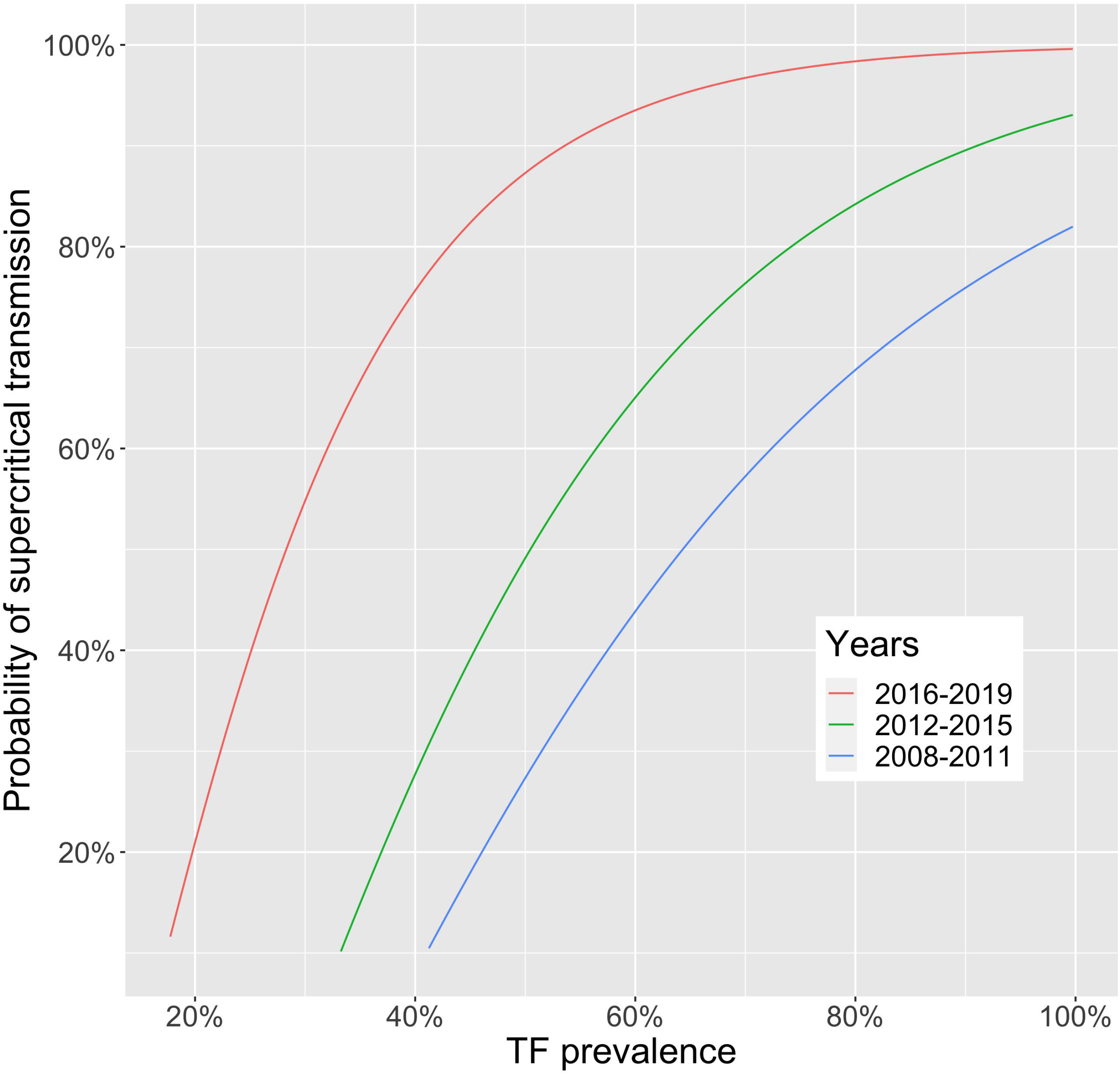
The probability that a district is a transmission-hotspot, as a function of a single TF prevalence measurement. Transmission-hotspots are defined as having a reproduction number >1 despite annual MDA. Districts with TF values lower than those shown correspond to regions where trachoma transmission is likely to be self-limited. 95% confidence intervals are shown in Figure S4.

## Discussion

When the GET2020 Alliance first met in 1997, there was not enough data to accurately assess trachoma prevalence, let alone forecast future disease burden. The more extensive surveillance in the past decade has permitted probabilistic forecasts for TF prevalence in 2020 and beyond. Progress can be seen in the increased proportion of districts with TF less than five percent. Clinical trials have shown that MDA contributes to this decline (over 770 million of azithromycin were distributed to over forty countries by 2019).^21,22^ Other strategies that may have helped include improved facial cleanliness and environmental improvement, although the efficacy of these interventions has yet to be demonstrated in clinical trials. Caution is needed in attributing all of the decrease to improved control though because surveillance of low endemicity districts likely increased substantially when the GTMP was initiated in 2012. An additional caveat is that after the initial rounds of baseline surveys conducted in the Global trachoma mapping Project, subsequent impact assessments have only occurred in districts in which there was MDA distribution. Districts without intervention do not get routinely surveyed, which would bias the observed average TF prevalence downwards.

The tendency of the TF distributions for recent years to approximate an exponential distribution is suggestive that the overall reproduction number for TF is less than one.^12,13,23,24^ Having a reproduction number less than one is a prerequisite for local elimination of disease. Although there may be other reasons why the TF distribution now appears exponential such as variable surveillance coverage, this finding provides additional reassurance that progress towards the WHO elimination goals has been made.

### Forecasts

Prior work has shown that forecasting the prevalence of TF in individual districts is challenging, particularly given the relative paucity of data.^25^ However, by combining data from all surveyed districts, the GET2020 database provides an opportunity to assess the overall prevalence of TF and the progress towards elimination. Of crucial importance, our forecasts are based on the assumption that intervention will continue at the current level of effort. To maintain current efforts, political will and financial support must remain favorably aligned.

The geographic correlation amongst the entries in the GET2020 database is unknown. Thus the effective number of entries in the database is likely lower than the actual number of entries, and this can serve to falsely accentuate the reliability of forecasts. In addition, while standardized diagnosis of trachoma has improved the reproducibility of trachoma prevalence surveys, sampling error is still expected.^26^ These biases are only partially compensated for by the confidence intervals we have determined via bootstrapping. As new data is made available, our forecasts will become falsifiable, and any misalignment would inspire a reevaluation of our model assumptions. For future research, spatial correlation may permit more accurate forecasts at regional levels.

### Transmission-hotspots

Our approach has been statistical and does not directly incorporate mechanistic details of trachoma transmission that predispose transmission-hotspots being refractory to annual MDA. Thus, while the overall burden of trachoma is clearly decreasing and many regions are reaching local elimination, the pathway to global control is not as deterministic as the forecasts may suggest.

By providing a probability that a TF prevalence measurement indicates a transmission-hotspot, our analysis highlights that some districts that have previously been identified as ‘hyperendemic’ may not necessarily be a transmission-hotspot. Rather, some variability in TF prevalence is expected, even when transmission is disappearing overall. As progress towards global control continues and districts with higher TF become increasingly likely to be transmission-hotspots, the need for enhanced surveillance and consideration of novel control strategies may become more targeted. In future work, it may become important to pursue within-district mapping for transmission-hotspots. Improved understanding of transmission dynamics will allow our proposed probabilistic relationship between trachoma prevalence and the probability of being a transmission-hotspot to be tested in a falsifiable manner.

## Conclusion

With the GET2020 Alliance, there have been substantial improvements in the reliability and coverage of trachoma surveillance. Surveillance data shows clear evidence of an increasing number of districts achieving local control with TF < 5%. Districts with TF of 5-20% are also likely to be representative of successful control programs, and eventual achievement of TF < 5% is expected. Districts with TF > 20% may represent transmission-hotspots that require more intensive treatment to achieve control. These results provide a quantitative basis for policy decisions (Table S1), and inspires several future areas of research. These types of analysis can help to direct resources away from areas where trachoma elimination appears imminent and towards challenging regions where additional interventions may be needed. Continued assessment of the global control of trachoma is needed so that the tremendous successes are not compromised when the decreased burden of disease may paradoxically reduce the political and economic will to continue elimination efforts.

## Supporting information

Supplementary references

## Data Availability

All code was conducted with R version 3.6.1 and is available on github (@proctor-ucsf/Trachoma-CID-2021-code). Due to the sensitive nature of data ownership amongst the Ministries of Health, the individual data are available by request.

https://github.com/proctor-ucsf/Trachoma-CID-2021-code

## Authors’ contributions

TML conceived the research. BFA, SB, AB, MSD, TDH, TML TCP, and JMP are part of the trachoma group in the NTD modeling consortium, whose ongoing group discussions inspired specific analyses. CEO contributed field expertise concerning model assumptions. MSD, PME, WWG, and PJH managed data. SB and CT performed analysis. SB wrote the first draft. All authors edited the manuscript and approved the final draft. The authors alone are responsible for the views expressed and they do not necessarily represent the views, decisions or policies of their affiliated institutions.

## Acknowledgements

The major contributors to the GET2020 database are the health ministries of trachoma-endemic countries worldwide. Generation of the data required examination of millions of people by thousands of field teams; field team trainers; supervisors; statisticians; epidemiologists; data analysts; project managers and many others. The roles of all of these stakeholders in generating the GET2020 database are recognized with grateful appreciation (Supplemental references).

