## Supplementary table for "Forecasting trachoma control and identifying transmission-hotspots"

| **Policy Relevant principle** | **Application to manuscript** | **Location of specific detail** |
| --- | --- | --- |
| **Stakeholder engagement** | Study inspired by the GET2020 Alliance. Assessment of trachoma control goals and identification of transmission-hotspots is of key importance to ongoing policy decisions. | Introduction |
| **Complete model documentation** | A complete description of our model is provided. Code is available in a GitHub repository | Methods    Github: @proctor-ucsf/Trachoma-CID-2021-code |
| **Complete description of data used** | Data is obtained from the GET2020 database, as maintained by ITI. | Methods |
| **Communicating uncertainty** | Assumptions of the model that lead to uncertainty in the results are described. Bootstrapping is used to generate confidence intervals | Methods/Discussion |
| **Testable model outcomes** | 1. Forecast of the distribution of TF prevalence for 2020-2030.  2. Probabilistic relationship  between TF prevalence and transmission-hotspots. | Results / Discussion |

***Table S1*** *- Table 2: Summary of Policy-Relevant Items for Reporting Models in Epidemiology of Neglected Tropical Diseases.**^35^*

*GET2020 Alliance =  World Health Organization Alliance for the Global Elimination of Trachoma by the year 2020; ITI = International Trachoma Initiative*
