## Supplementary text for "Forecasting trachoma control and identifying transmission-hotspots"

**Data**

The GET2020 database is managed by the International Trachoma Initiative (ITI) in collaboration with the World Health Organization (WHO) on behalf of the GET2020 Alliance. Some data such as district-level prevalence is publically available, but ownership of the data resides with the Ministries of Health worldwide.^1^

As part of the effort to eliminate trachoma, the quality and coverage of trachoma surveillance has increased substantially in recent years.^2^ In a typical trachoma prevalence survey, 1000+ children (aged one to nine years) from 20-30 villages are examined for TF. Health ministries contribute to the database by submitting survey results to WHO and ITI; key partners have included the Global Trachoma Mapping Project (GTMP) which supported health ministries to examine 2.6 million people in 29 countries from just 2012-2016^3,4^ and Tropical Data, which has provided analogous support since 2016. While it is not a direct marker of infection, TF prevalence is a useful tool for monitoring the burden of trachoma because it is relatively easy to measure and reflects the overall burden of ocular disease in a district. In fact, one of the requirements for validation by WHO for control is that the TF prevalence in 1-9 year olds in a formerly endemic district must remain less than five percent for at least two years in the absence of ongoing antibiotic MDA.

Because the GET2020 database covers a huge geographic area, not all districts in all endemic countries are surveyed every year. Following WHO guidance, trachoma prevalence surveys are conducted in a predictable pattern: 1) Districts suspected of having endemic trachoma may conduct a baseline survey, 2) Districts having a substantial gap in time (typically ten years) from an initial baseline survey and no intervention may conduct a new baseline survey, to re-evaluate the need for MDA, 3) Districts that have initiated azithromycin MDA will conduct an impact survey six to twelve months after all MDA cycles have been completed, 4) Districts will conduct a surveillance survey at least two years after an impact survey reported TF prevalence <5% and MDA was stopped to assess any resurgence.

Prior to 2012, prevalence surveys were largely conducted by health ministries in partnership with non-governmental organizations. Between 2012 and 2016, the GTMP helped to broaden survey coverage and standardize survey systems and methodologies; Tropical Data now continues that work using the same methodologies. ITI facilitates donation of antibiotics based on the results of prevalence surveys. If TF prevalence is greater than or equal to 5, 10 or 30 percent, ITI provides azithromycin for one, three or five annual rounds of antibiotics, respectively. Districts for which TF prevalence is less than five percent do not qualify for antibiotic MDA.

**Data cleaning**

Our model was trained with all TF prevalence measurement entries in the GET2020 database for 2004-2019, with two exceptions. First, TF prevalence measurements of less than 0.5% were not included, as these may not have needed to be surveyed. The vast majority of districts worldwide fall into this category, but only a small minority are surveyed. Second, to reduce the bias of introducing surveys obtained outside the scope of WHO guidelines for trachoma control, we only used survey data that was clearly labeled as baseline, impact or surveillance. Third, when there were identical TF prevalence entries for multiple districts in the same geographic region for the same survey year, we retained one randomly selected datum. Our rationale was that sometimes TF survey data were not resolved to the district level and these instances showed up in the database as multiple entries.

**Forecasting model**

Our use of a susceptible-infected-susceptible model assumes that among children aged 1-9 years within a single district, there is uniform mixing, each child has the same intrinsic susceptibility, and there is no variability in bacterial load or transmission potential among those infected. Infection occurs in proportion to the product of those who are infected and those who are not infected. Those who are infected become susceptible again at a constant rate, without any residual immunity. Further, our model assumes that the presence of TF acts as a reasonable surrogate for infection, even though it actually represents the inflammatory sequelae of infection and can persist long after infection clears. The advantage of using the susceptible-infected-susceptible model is that it implies that if trachoma is disappearing over time, the cross-sectional distribution of district-level TF prevalence for each year follows an exponential distribution (i.e. the discrete counterpart to an exponential distribution).^5,6^ Although these assumptions are not entirely consistent with our nuanced understanding of trachoma biology and population structure, the resulting model has been shown to parsimoniously reflect epidemiological observations in a statistically robust manner.^7^

Our exponential distribution model for the observed TF prevalence distribution for each yar is equivalent to a constant proportional decrease in the reproduction number each year. To calibrate this model with the observed data, the TF prevalence probability distribution model for each year was truncated to satisfy 0.5% < TF < 100% and renormalized so that the integral of the probability density is the unity. In a secondary analysis, we also explored alternate models based on the corresponding truncated versions of the beta distribution (parameterized by two shape parameters), gamma distribution (parameterized by a shape and rate parameter), and Lomax distribution (parameterized by a shape and scale parameter). The parameters in the alternate models were all assumed to follow an exponential curve over time.

The likelihood of each datum was computed by determining the model’s probability density function for the corresponding year and TF prevalence value. Maximization of the likelihood determined the two coefficients describing the intercept and decay of each model parameter.

The 95% confidence interval for our forecast was obtained by bootstrapping the GET2020 database with replacement. In each bootstrap replicate, we resampled the list of countries with replacement, built a dataset by combining the data for each randomly chosen country (sometimes with duplication), and re-fit the forecast model with the district-level data for the chosen countries. The distribution created by the ensemble of bootstrap forecasts determined the confidence intervals. Bootstrapping was performed at the country level, in order to minimize correlation within the data.

**Reproduction number estimate**

Our probabilistic forecasts allow the opportunity to estimate the effective reproduction number, *R*, for trachoma. This number represents the average number of new TF cases that are caused by a single TF case under current interventions. In a susceptible-infected-susceptible model in which the proportion of individuals in the population is slowly changing, the proportion of infected satisfies,^8^

$I(t) = e^{\gamma(R-1)t}$.

Here, *t*, is time and, $\gamma$, is the rate at which infected individuals recover. Based on prior analyses, we estimate $\gamma$to be 2/year, corresponding to a TF duration of six months.^9^ The log of the ratio of TF for two successive years is then $2(R-1)$, which can be solved for R.

**Transmission-hotspots**

A common approach to trachoma epidemiology is to set specific thresholds of TF prevalence to classify the degree of transmission in a particular district. For example, districts may be classified as being hypo-endemic for TF of 5-9%, mesoendemic for TF of 10-29% or hyperendemic for TF >= 30%.^29,30^ However, this classification does not correlate directly with measures of transmissibility because it ignores the stochasticity that is inherent in trachoma transmission. This stochasticity implies that there can be a long tail to the TF distribution even for districts that are progressing towards elimination. Thus, a single TF measurement is typically insufficient to determine whether a particular value of TF corresponds to a transmission-hotspot district. Instead, a TF measurement can be used to assign a probability that a district is a transmission-hotspot.

By modeling the district-level prevalence as an exponential distribution, our forecast assumes that the prevalence of trachoma is decreasing throughout the world. In reality, there remains some transmission-hotspots where control has been challenging to achieve and other districts in which intervention has not started. To quantify the proportion of districts that are transmission-hotspots and how this proportion changed over time, we divided the GET2020 database into four year intervals. For each interval, we fit truncated exponential and gamma distributions by maximum likelihood. As above, the distributions were truncated and renormalized to satisfy 0.5% < TF < 100%. The gamma distribution permits a heavier tail to the TF distribution such as may occur when transmission-hotspot districts do not respond to MDA.

For a specific observed value of TF, we estimate the probability that a district is a transmission-hotspot as the proportion of the truncated gamma distribution density that is greater than the truncated exponential distribution. The density of the truncated exponential distribution forms a null model since it is what we would expect to observe by chance if all districts were subcritical (e.g. reproduction number < 1). We only report transmission-hotspot probabilities that are greater than 10%, because this method is unreliable for low values of TF where districts are likely to be subcritical (i.e. mathematically, the gamma distribution density becomes less than the exponential distribution density).

**Reproducibility**

All code was conducted with R version 3.6.1 and is available on github (@proctor-ucsf/Trachoma-CID-2021-code). Due to the sensitive nature of data ownership amongst the Ministries of Health, the individual data are available by request.

11.	Borlase, Anna. Modelling trachoma post 2020: Opportunities for mitigating the impact of COVID-19 and accelerating progress towards elimination. *Transactions of the Royal Society of Tropical Medicine and Hygiene*. ((In review)).
