## Supplementary figures for "Forecasting trachoma control and identifying transmission-hotspots"

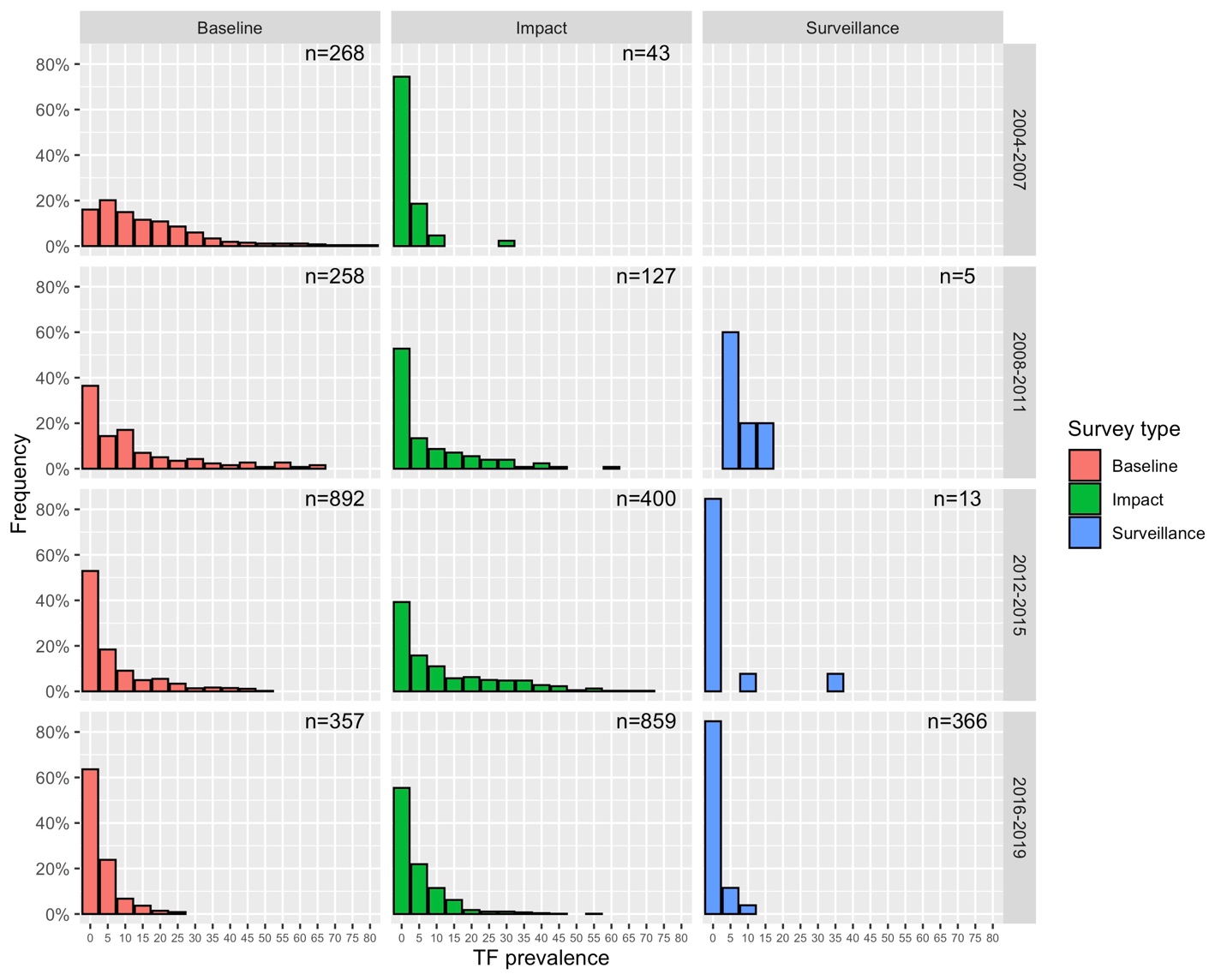

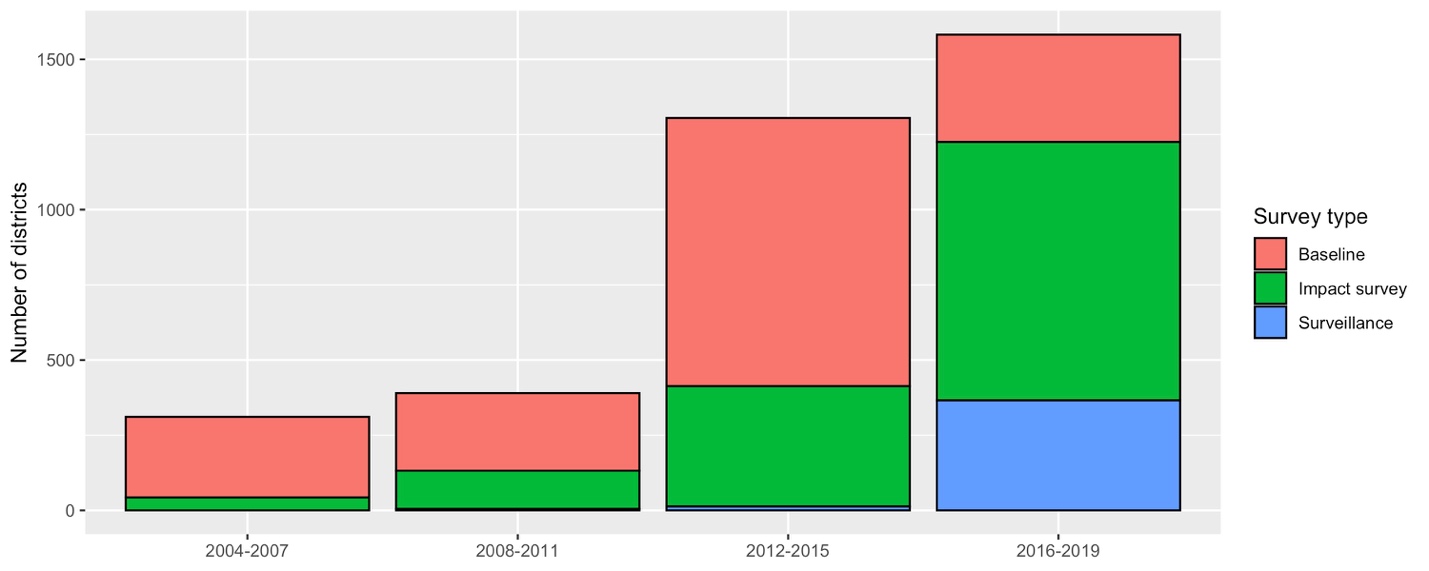

Figure S1: (Top) *Distribution of district-level prevalence of TF as recorded by the GET 2020 alliance* stratified by survey type and four year intervals. (Bottom) Overall number of surveys of each type, grouped by the four year intervals.

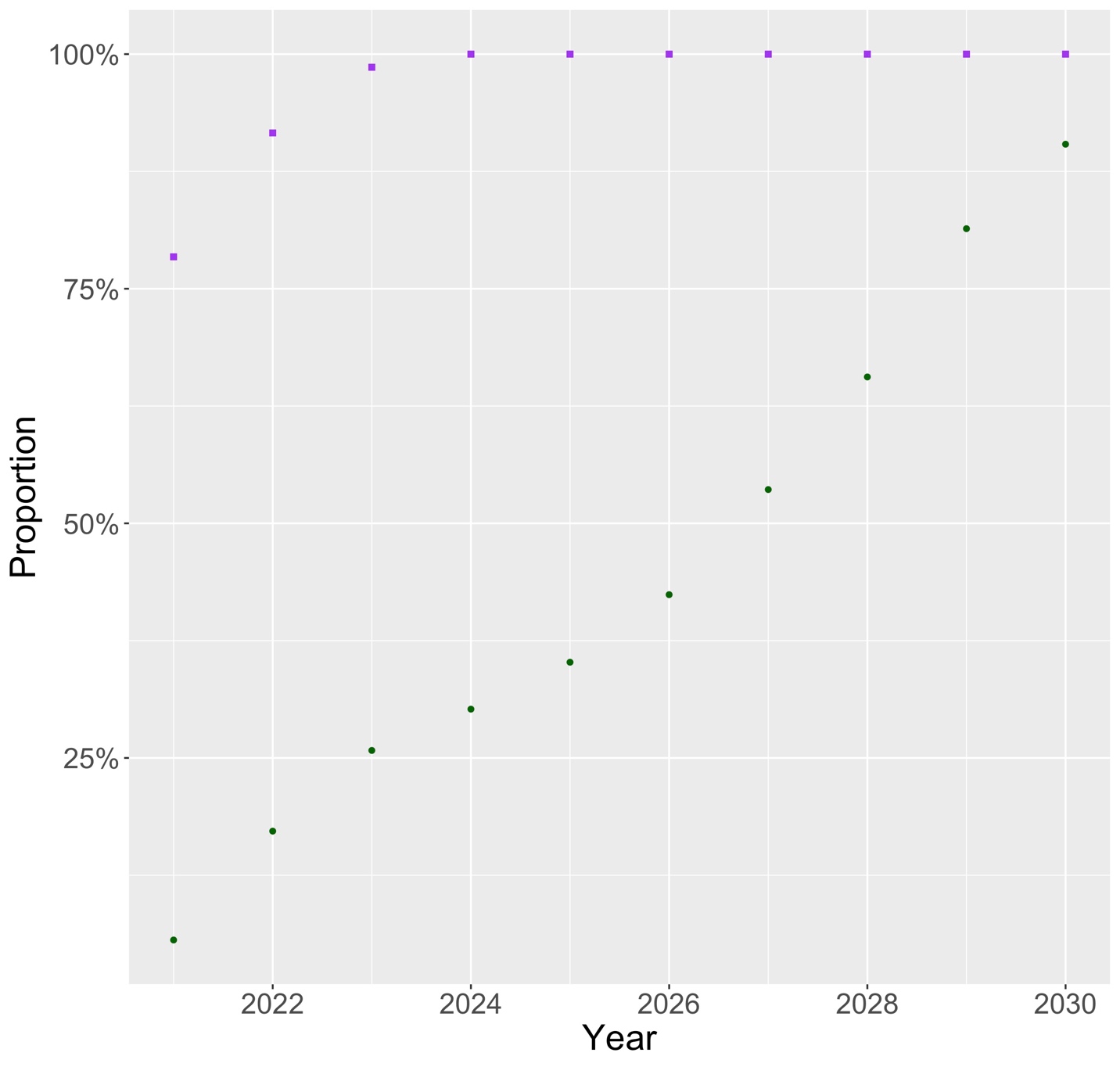

Figure S2: Percentage of bootstrap stimulations that have a mean TF prevalence < 5% (purple squares) and have over 90% of districts with TF < 5% (green circles).

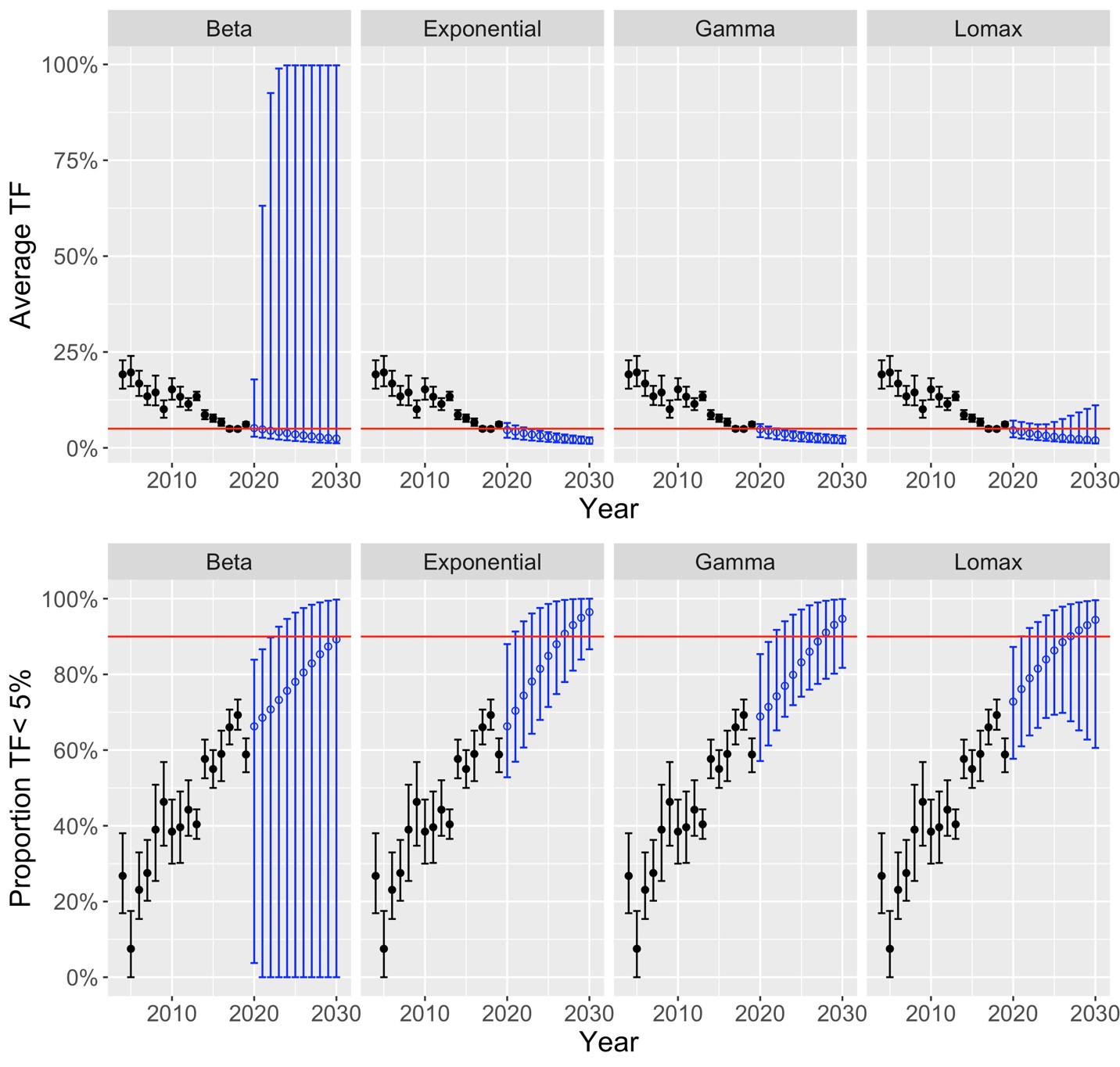

Figure S3: Mean TF prevalence of all surveyed districts (top) and percent of surveyed districts with TF less than 5 percent (bottom). Each vertical column corresponds to a different distribution used for modeling the TF prevalence in any given year. The format of the graphs is identical to that seen if Figure 2 and the panels corresponding to the exponential distribution are a replicate of Figure 2. Additional methodological details concerning the probabilistic forecasts can be found in the Methods Section of the main text.

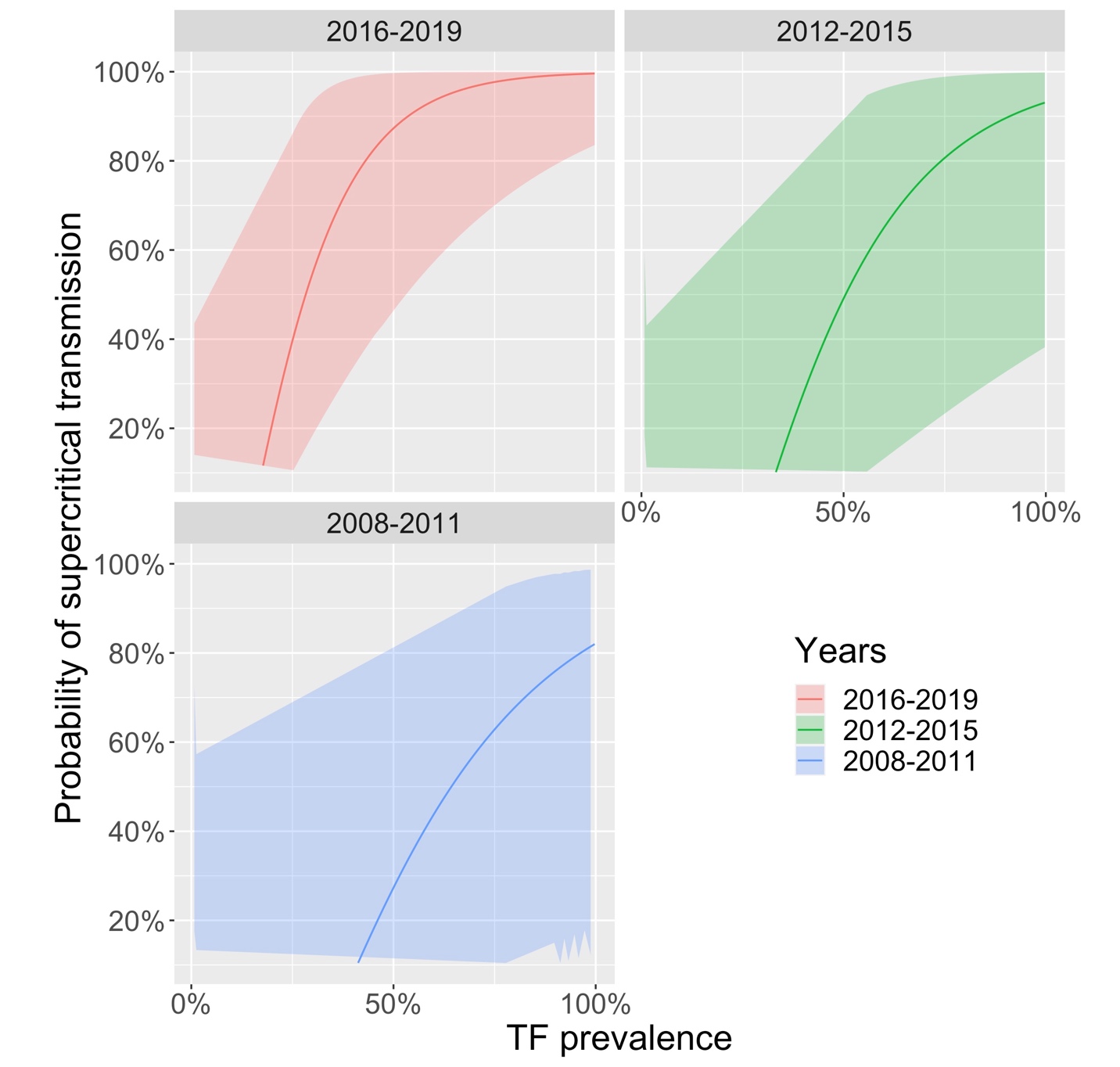

Figure S4: The probability that a district is a transmission-hotspot, as a function of a single TF prevalence measurement. Each panel represents a four-year period. Solid lines are replicates of Figure 3 of the main text and show results for the observed data. The shaded regions are 95% confidence intervals as determined by replicating the analysis on an ensemble of country-level bootstraps.
